# Olfactory Dysfunction in Patients with Multiple Sclerosis; A Systematic Review and Meta-Analysis

**DOI:** 10.1101/2021.06.16.21259033

**Authors:** Omid Mirmosayyeb, Narges Ebrahimi, Mahdi Barzegar, Alireza Afshari-Safavi, Sara Bagherieh, Vahid Shaygannejad

## Abstract

**Objectives:** Olfactory dysfunction is a major comorbidity observed in patients with multiple sclerosis, yet different prevalence rates are reported for it. Therefore, we have designed this systematic review to estimate the pooled prevalence of olfactory dysfunction in patients with MS. To our knowledge, this is the first systematic review and meta-analysis on the prevalence of olfactory dysfunction in MS patients.

**Method:** We searched PubMed, Scopus, EMBASE, Web of Science, ProQuest, and gray literature including references from the identified studies, review studies, and conference abstracts which were published up to January 2021. Articles that were relevant to our topic and could provide information regarding the prevalence of olfactory dysfunction, or the scores of smell threshold, discrimination, or identification (TDI scores) among MS patients and healthy individuals were included; however, articles published before 1990 and after the end of 2020 were excluded.

**Results:** The literature search found 1630 articles. After eliminating duplicates, 897 articles remained. two abstract conference papers were included for final analysis. A total of 1099 MS cases and 299 MS patients with olfactory dysfunction were included in the analysis. The pooled prevalence of olfactory dysfunction in the included studies was 27.2%. (95% CI: [19.7%, 35.4%]) Also, the overall TDI score in MS patients was lower than that in the control group (SMD=-1.00; 95% CI: [−1.44, −0.56]), and the level of Threshold (SMD= −0.47; 95% CI: [−0.75, −0.19]), Discrimination (SMD=-0.53; 95% CI: [−0.96, −0.10]), and Identification (SMD=-1.02; 95% CI: [−1.36, −0.68]) per se were lower in MS compared with control respectively.

**Conclusion:** The results of this systematic review shows that the prevalence of olfactory dysfunction in MS patients is high and more attention needs to be drawn to this aspect of MS.

## Introduction

Olfaction is one of the most overlooked sensations of the human being. It is the means to the perception of smell, recognizing imminent dangers, and even storing memories and emotions.(1–3) The actual mechanism by which olfaction is mediated is still not completely clear to this date; however, data suggest that an interaction between odor receptors and odor molecules initiate the process, leading to the production of olfactory signals which travel through the olfactory nerves to the Central Nervous System (CNS). This path is then followed by the storage of the smell as memories within the CNS for faster, more appropriate, and more reliable reactions in case of future encounters.(4,5)

Multiple Sclerosis is an autoimmune disease, characterized by demyelination of CNS tissue. MS is a lifelong condition that can affect the brain and spinal cord, leading to a wide range of symptoms, including problems with vision, motor control, cognitive abilities, balance, and sensation. One of the most vulnerable of the sensations is olfaction as it is in direct contact with patients’ physical, behavioral, and cognitive state. Olfaction is shown to be prone to impairment in three main aspects, including threshold, discrimination, and identification. Olfactory dysfunctions are reported as one the most common manifestations in the initial stages of certain CNS diseases, including Alzheimer’s disease and Parkinson’s disease. Moreover, numerous studies have pointed out the presumable connection between olfactory disturbances and not only neurodegenerative diseases but also autoimmune ones, such as Multiple Sclerosis (MS).(2,6) Several factors can contribute to the olfactory dysfunction MS patients occasionally report, some of which include persistent inflammation within the CNS, demyelination of olfactory bulbs, and the burden of plaque in brain areas associated with the olfactory system. Previous original articles have shown that the prevalence of olfactory disruption is higher among MS patients than healthy individuals.(2,6,7)

Previous studies have provided conflicting evidence on determining the specific aspect of olfaction that suffers the most among MS patients, such aspects include Threshold, Discrimination, and Identification (TDI) dysfunction. Such studies also lack coherence regarding the prevalence they report, with numbers ranging from 20% to 40%.(8,9) It is crucial to study olfactory dysfunction as it plays a major role in diminishing one’s quality of life,(10) and also because there is growing evidence that the degree to which MS patients present with olfactory problems can be used as a potential prognostic factor.(11) On the other hand, the prevalence of olfactory disturbance among MS patients has been reported in various studies, among different sample sizes with different MS subtypes. Consequently, we designed this systematic review and meta-analysis to estimate the pooled prevalence of olfactory dysfunction among MS patients. The aims of this study are to: 1-Estimate the pooled prevalence of olfactory dysfunction among MS patients and 2-Compare the TDI score among MS patients and healthy individuals.

## Methods

### Literature search

We conducted a systematic computerized search using four data banks: PubMed (MedLine), Scopus, web of science, and Embase (via Elsevier), and ProQuest. We also searched the gray literature including references from the identified studies, reviews studies, and conference abstracts which were published up to January 2021.

### Inclusion criteria

Studies reporting the prevalence of olfactory dysfunction or the scores of Threshold, Discrimination, and Identification (TDI) among MS participants regardless of the diagnostic method with a sample size of over at least 10 patients were included.

Nevertheless, case reports and case series articles, articles that were written in any language other than English, and any published studies before 1990 and after the end of 2020 were excluded.

### Data search and extraction

We conducted a systematic computerized search using four data banks: PubMed (Medline), Scopus, web of science, Embase, and ProQuest. We also searched the gray literature including references from the identified studies, reviews studies, and conference abstracts which were published up to January 2021.

We used Mesh terms and text words to generate a syntax that included two components.

We used Mesh terms to generate a syntax that included two components.

“Olfaction Disorder,” “Smell Disorder”, “smell dysfunction”, “olfactory agnosia”, “agnosias for smell”, “dysfunction AND smell”, “olfactory impairment”, “impairment AND olfactory”, “sense of smell”, “smell sense”, “loss of smell”, “smell loss”, “Cacosmia”, “Dysosmia”, “Anosmia”, “paraosmia”, “hyposmia”, “agnosias”, and “agnosia AND olfactory” were the keywords we used to describe olfactory dysfunction; and also “multiple sclerosis”, “MS”, “disseminated sclerosis”, “Sclerosis AND multiple”, “sclerosis AND disseminated”, “acute fulminating”, and “acute fulminating” were the keywords we used to identify the other search component. Additionally, we customized our search syntax (query) for each data bank.

Two researchers (NE and SB) independently screened the articles. Any disagreement between the aforementioned researchers would be addressed by the senior researcher of the team (OM). The data extraction table included first author, region of study, date of publication, type of study, sample size of case and control group, and the demographic variables for case and control such as sex and mean of age. Other variables that we collected in our table included the exact name of the olfaction screening test, MS subtype, disease duration, EDSS score, number of hyposmia and anosmia in both case and control, plus the mean and standard deviation of the Threshold, Discrimination, and Identification (TDI) scores if applicable. Had any of the included articles used over one diagnostic method, each different methods would have been mentioned in a separate row of the table with its respective data.

Furthermore, the olfaction diagnosis extraction form consisted of the total number of patients with olfactory dysfunction, hyposmia, anosmia, microsmia, and identification, threshold, and discrimination dysfunction.

All these variables were extracted from both MS and control group. In the present study, the control group represents healthy individuals without any neurologic disorders and/or diseases.

### Statistical analysis

A statistical test for between study heterogeneity was performed by I-square (I^2^) and Cochran’s chi-square test. If evidence of heterogeneity was observed, a random effect model was used. Forest plot was conducted to demonstrate the prevalence of olfactory in each study and the pooled estimate of prevalence with their 95% confidence intervals (95% CI). Subgroup analysis was performed by sample size (≤ 50 and > 50), publication year (≤ 2010 and > 2010), EDSS (≤ 3 and > 3) and disease duration (≤ 10 and > 10). Publication bias was assessed using funnel plot of logit transformed prevalence and Egger’s test. The Trim and fill approach was applied to obtain an adjusted effect size, when evidence of publication bias was observed. level of statistical significance was considered to be less than 0.05. All statistical analyses were done using Stata 14 software (Stata Corporation, College Station, Texas, USA).

## Results

The literature search found 1630 articles. After eliminating duplicates, 897 articles remained. Two abstract conference papers were included for final analysis. A total of 1099 MS cases and 299 MS patients with olfactory dysfunction were included in the analysis. For those included articles that had used more than one diagnostic method, we assigned separate rows for each of their methods. Hence, some articles have been mentioned more than once in Table 1 pertaining to the article’s different diagnostic means.

**Table 1.**
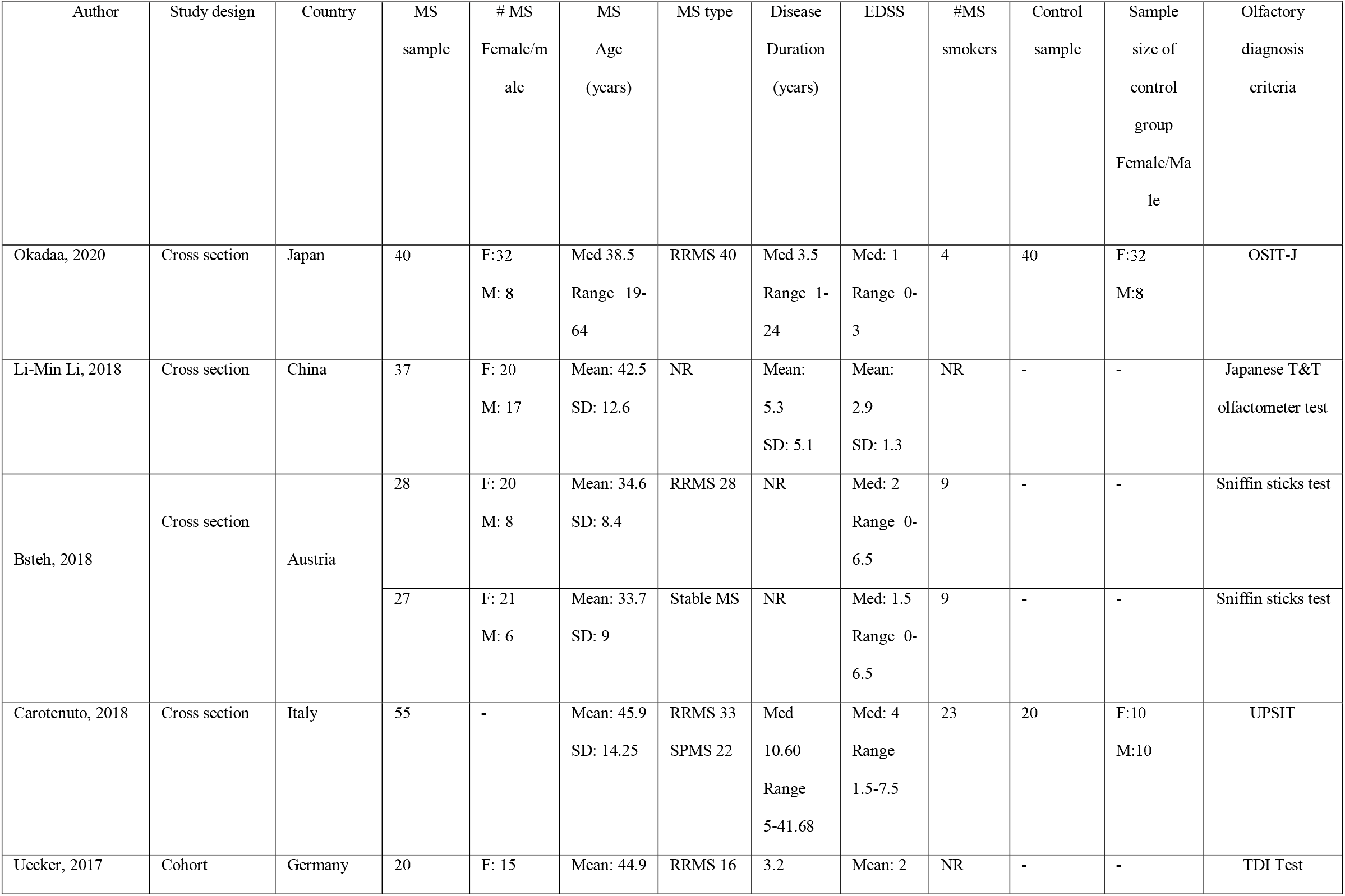
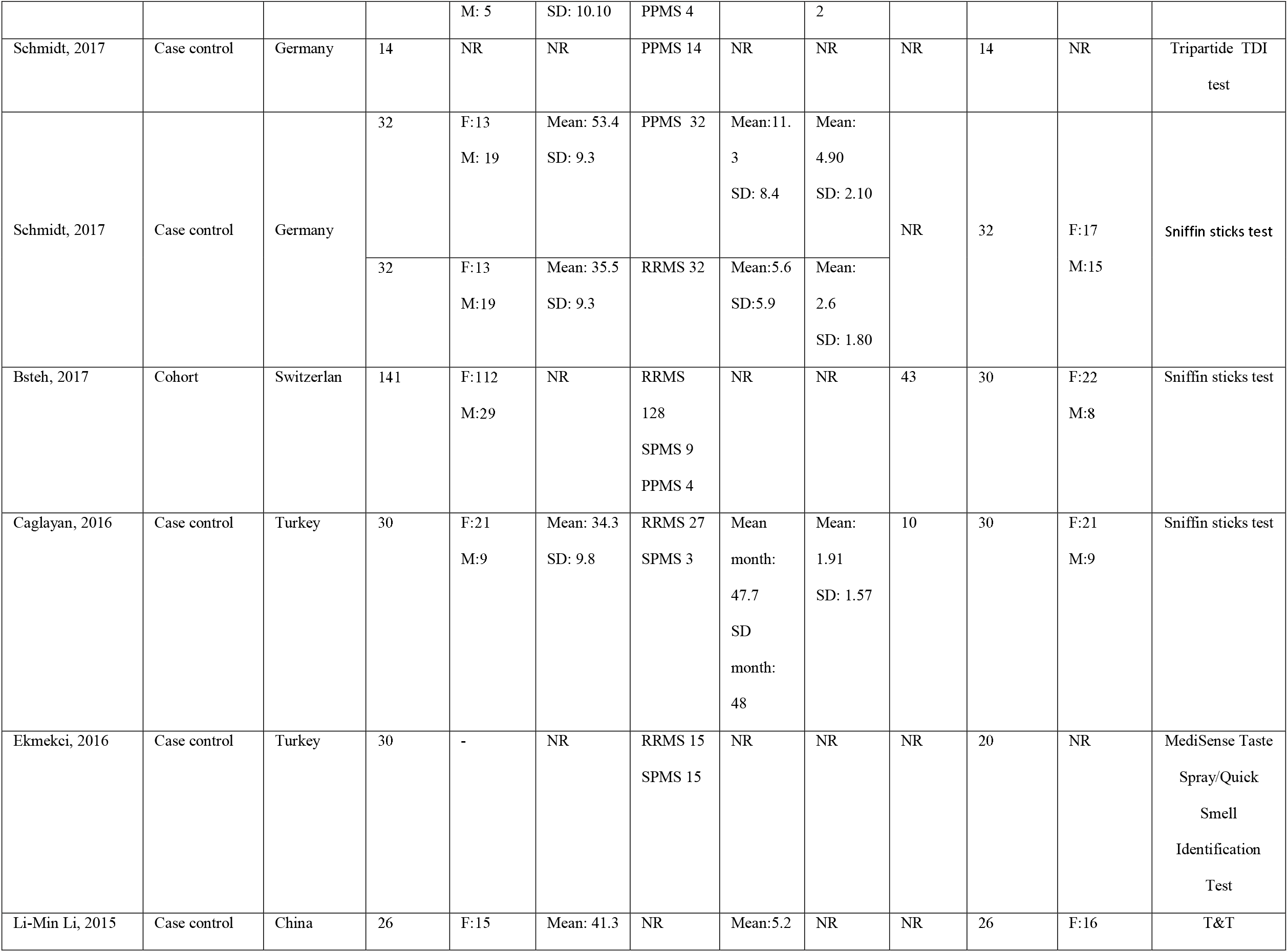
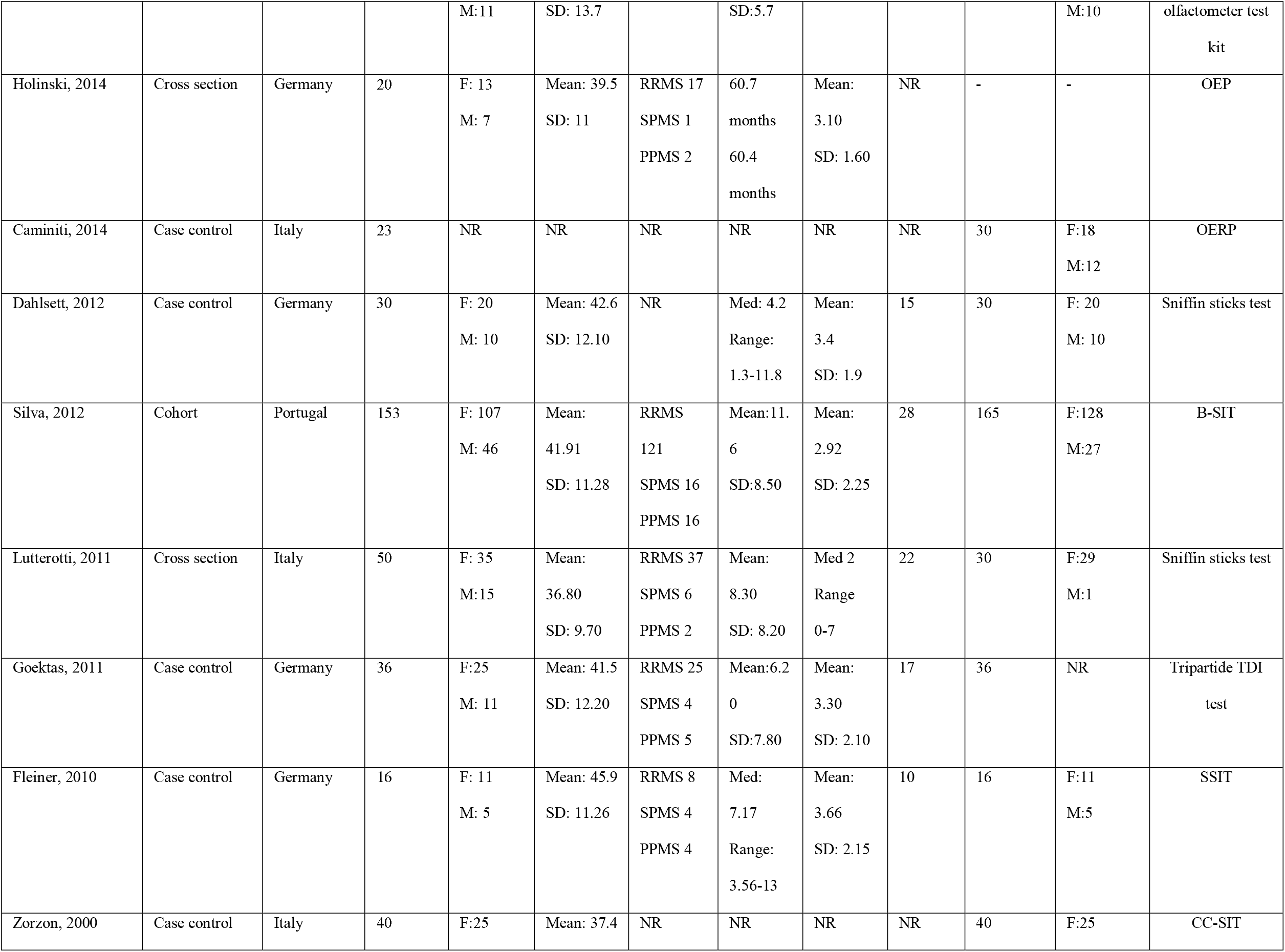
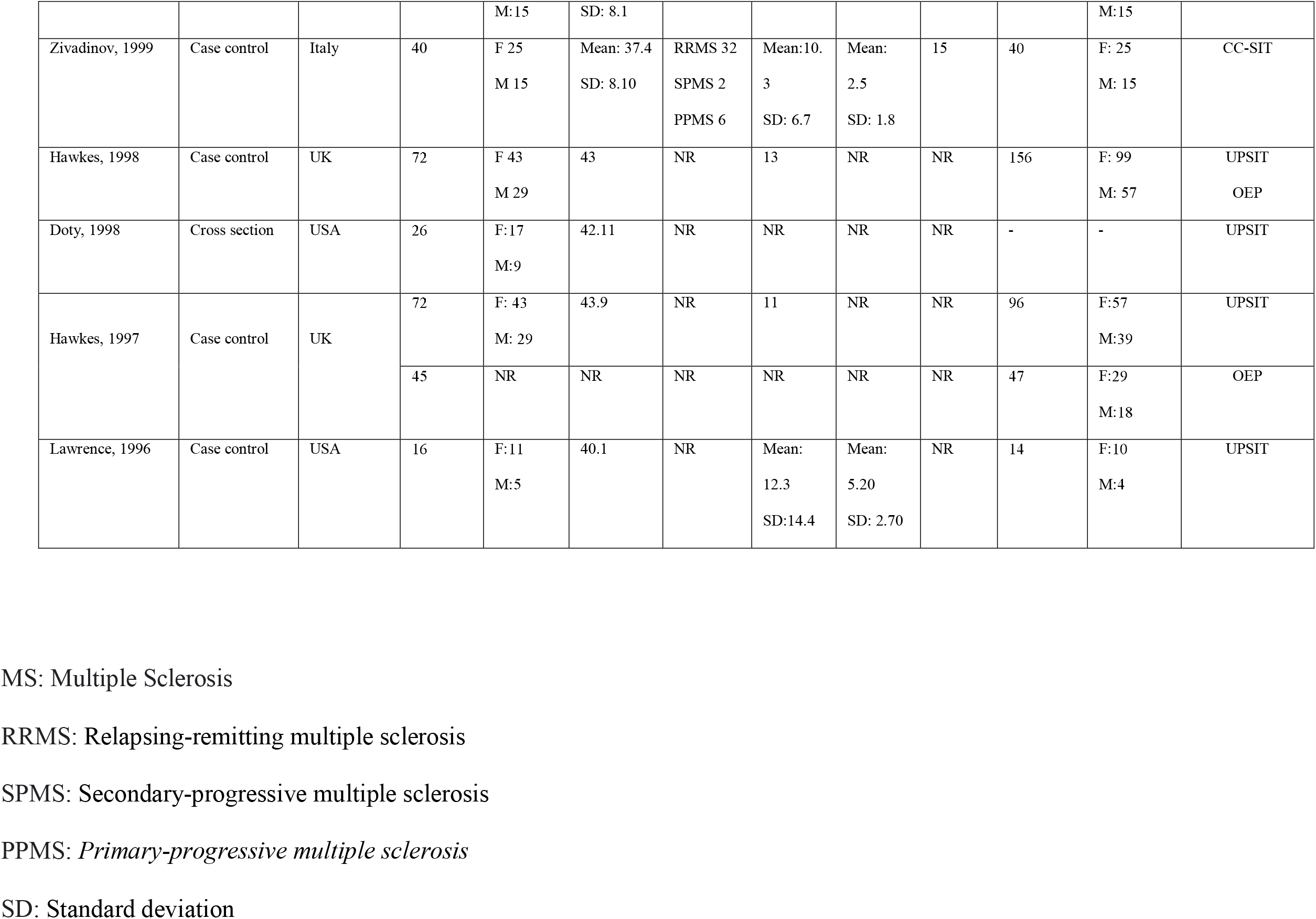
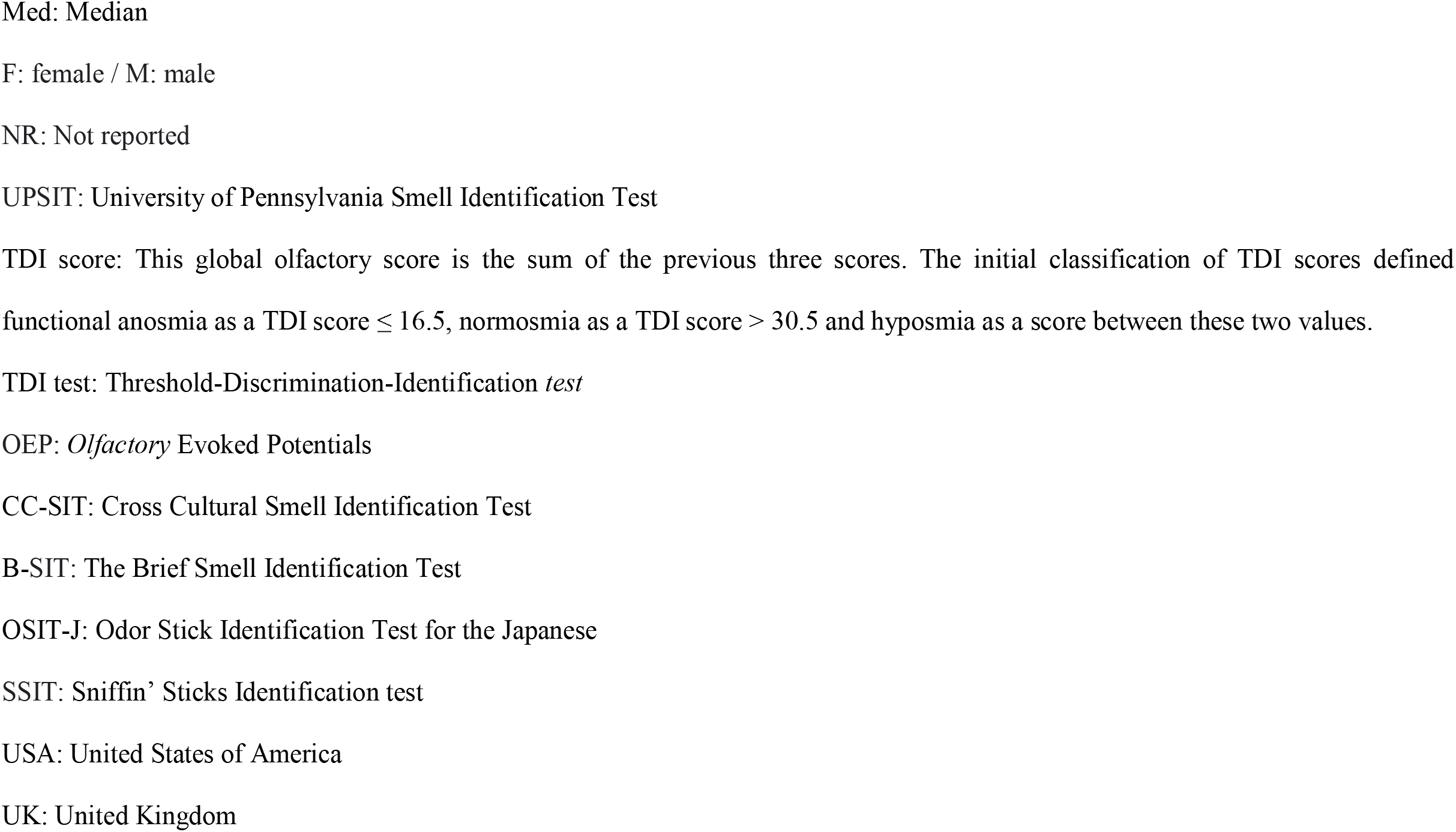
Basic characteristics of the included studies:

The pooled prevalence of olfactory dysfunction in the included studies was 27.2% (95% CI: [19.7%, 35.4%]) *Figure 1* As such, Bsteh (a) and (b), Hawkes (a) and (b), Schmidt (a), (b), (c), and (d), and Dahlsett (a) pertain to the different diagnostic methods that the aforementioned authors have used in their studies.

**Figure 1.**
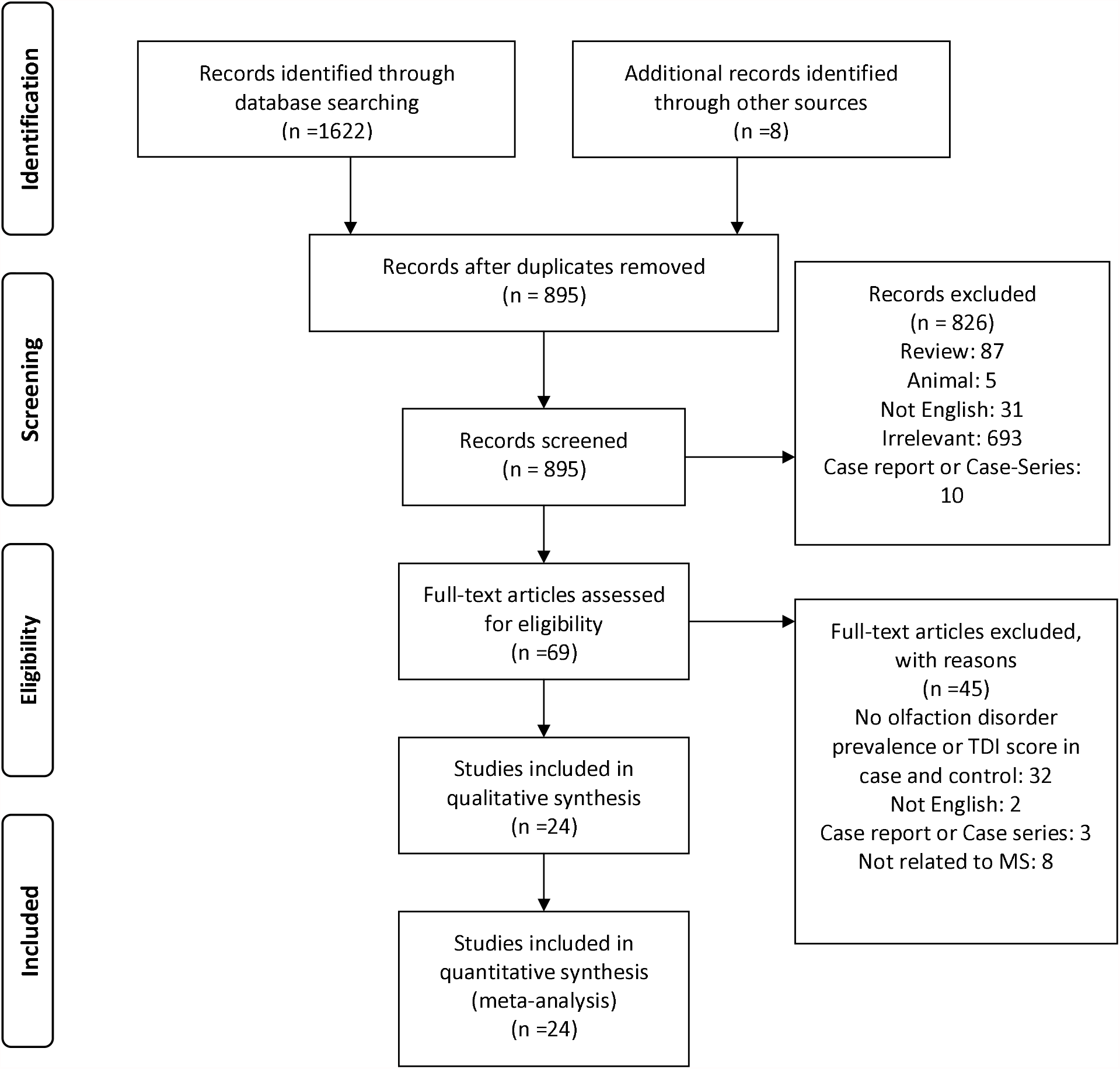
Flow diagram summarizing the selection of eligible studies.

### Critical appraisal

The quality of all the included articles was assessed using the Joanna Briggs Institute (JBI) critical appraisal checklist. The JBI checklist is the preferred tool for measuring the quality of descriptive studies reporting prevalence data and has a system of ranking articles based on the number of “YES” answers they earn according to its questions. The number of “YES” answers an article can earn ranges between 0 to 9.(12) Using this checklist, … Of the included studies earned less than 4 “YES” answers, … studies earned between 4 to 6 “YES” answers, and … studies earned more than 6 “YES” answers. *Figure 2, Supplementary 1, and Supplementary 2*

**Figure 2.**
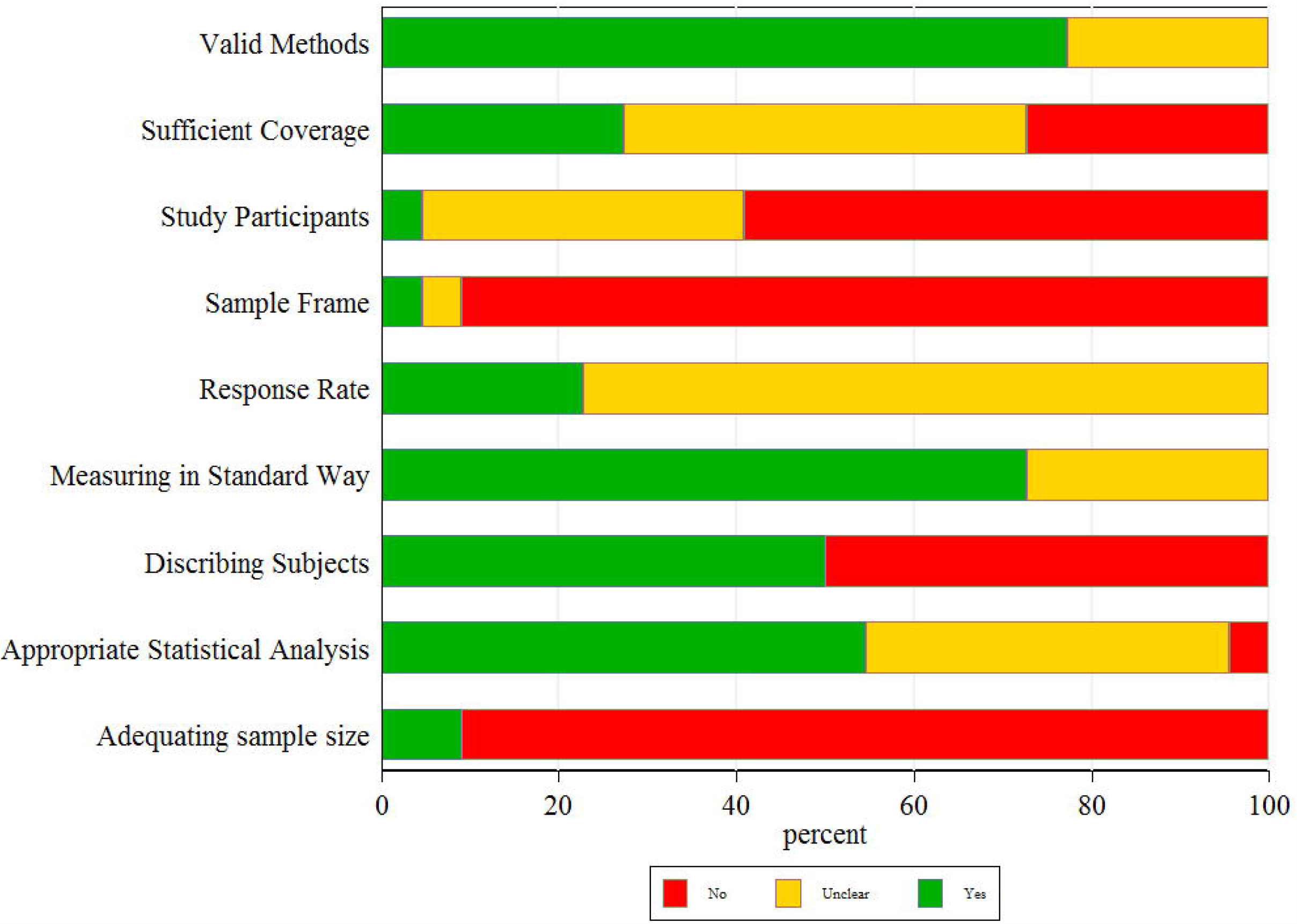
Quality assessment of the included studies based on the JBI checklist.

*Supplementary 1 Table of quality assessment of the included studies using the JBI checklist*.

*Supplementary 2 Figure of quality assessment of the included studies using the JBI checklist*.

### Prevalence estimates

The pooled prevalence of olfactory among patients with MS was 27.2% (95% CI: [19.7%, 35.4%]) *Figure 3* with a high level of heterogeneity (I^2^=87.4%; p<0.001) The prevalence estimates ranged from 0% observed in Austria population to 69.6% for Italy. The funnel plot *Figure 4* showed no evidence of publication bias, which was statistically supported by Egger’s regression test (Bias= 0.099; p=0.964).

**Figure 3.**
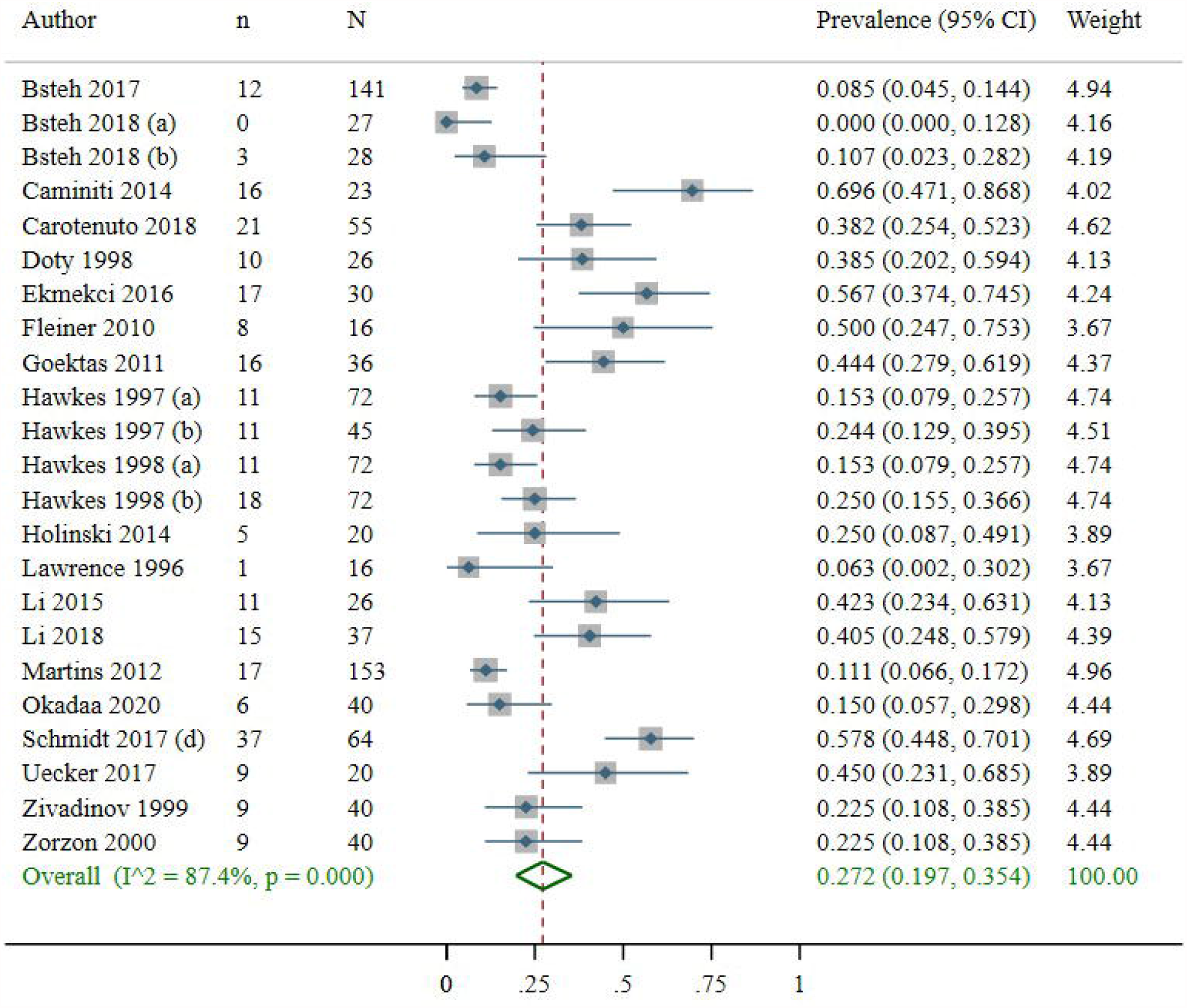
The pooled prevalence of olfactory among patients with MS.

**Figure 4.**
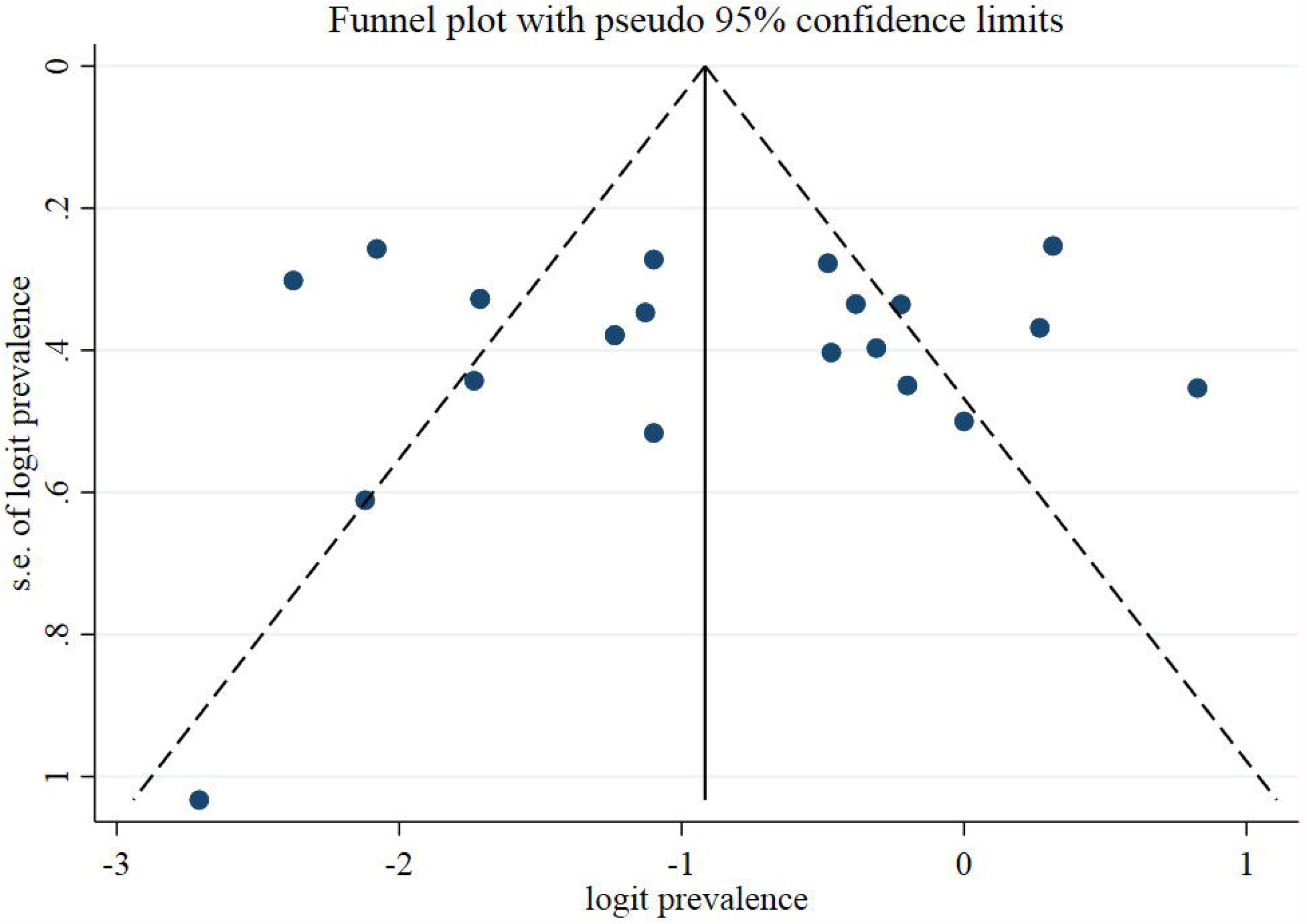
The funnel plot showing no evidence of publication bias, statistically supported by Egger’s regression test.

### Subgroup analysis

The results of subgroup analysis were shown in Table 2 by sample size, publication year, EDSS and disease duration. The pooled prevalence of olfactory was higher in studies with a mean EDSS more than 3 compared to those EDSS lower than 3 (32.6% vs. 15.9%, p=0.048). However, prevalence of olfactory was not significantly different in terms of sample size (15% vs. 22.6%, p=0.246), publication year (22.3% vs. 30.2%, p=0.373) and disease duration (36.1% vs. 18.7%, p=0.059).

**Table 2:** Subgroup analysis of pooled prevalence of olfactory.

| **Table 2: Subgroup analysis of pooled prevalence of olfactory** |  |  |  |  |  |  |
| --- | --- | --- | --- | --- | --- | --- |
| Subgroup by | | No. of studies | Total sample size | Pooled prevalence (95% CI) | Heterogeneity $I^2$ | p-value |
| Sample size | $\leq 50$ | 16 | 470 | 15% (5.7%-29.8%) | 81.47 | 0.246 |
| | $> 50$ | 7 | 629 | 22.6% (11.7%-35.6%) | 92.17 | |
| publication year | $\leq 2010$ | 9 | 399 | 22.3% (16.3%-28.9%) | 50.97 | 0.373 |
| | $> 2010$ | 14 | 700 | 30.2% (18.3%-43.4%) | 91.77 | |
| EDSS | $\leq 3$ | 8 | 486 | 15.9% (8%-25.6%) | 82.94 | 0.048 |
| | $> 3$ | 5 | 143 | 32.6% (19.4%-47.3%) | 64.89 | |
| Disease duration | $\leq 10$ | 7 | 195 | 36.1% (25.8%-47.1%) | 56.65 | 0.059 |
| | $> 10$ | 7 | 480 | 18.7% (12%-26.4%) | 72.75 | |

### Publication bias

Eight studies reported TDI, Threshold, Discrimination and Identification (220 controls and 240 cases) *Figure 5* The overall TDI score in MS patients was lower than that in the control group (SMD=-1.00; 95% CI: [−1.44, −0.56]). Also, overall level of Threshold (SMD= −0.47; 95% CI: [−0.75, −0.19]), Discrimination (SMD=-0.53; 95% CI: [−0.96, −0.10]) and Identification (SMD=-1.02; 95% CI: [−1.36, −0.68]) were lower in MS compared with control, respectively *Figure 6, Figure 7, and Figure 8* Between study heterogeneity was observed in all 4 indices, however we did not find any evidence of publication bias Table 3 and there was no need for additional studies.

**Table 3:** Level of Heterogeneity and Publication bias amongst the included studies

|  |  |  |  |  |  |
| --- | --- | --- | --- | --- | --- |
|  | Heterogeneity |  |  | Publication bias |  |
|  | Cochran's Q | I<sup>2</sup> | p | Egger's value | p |
| TDI | 33.33 | 79% | <0.001 | -6.522 | 0.076 |
| Threshold | 15.35 | 54.4% | 0.032 | -0.733 | 0.844 |
| Discrimination | 34.30 | 79.6% | <0.001 | -5.934 | 0.177 |
| Identification | 20.10 | 65.2% | 0.005 | -4.919 | 0.140 |

**Figure 5.**
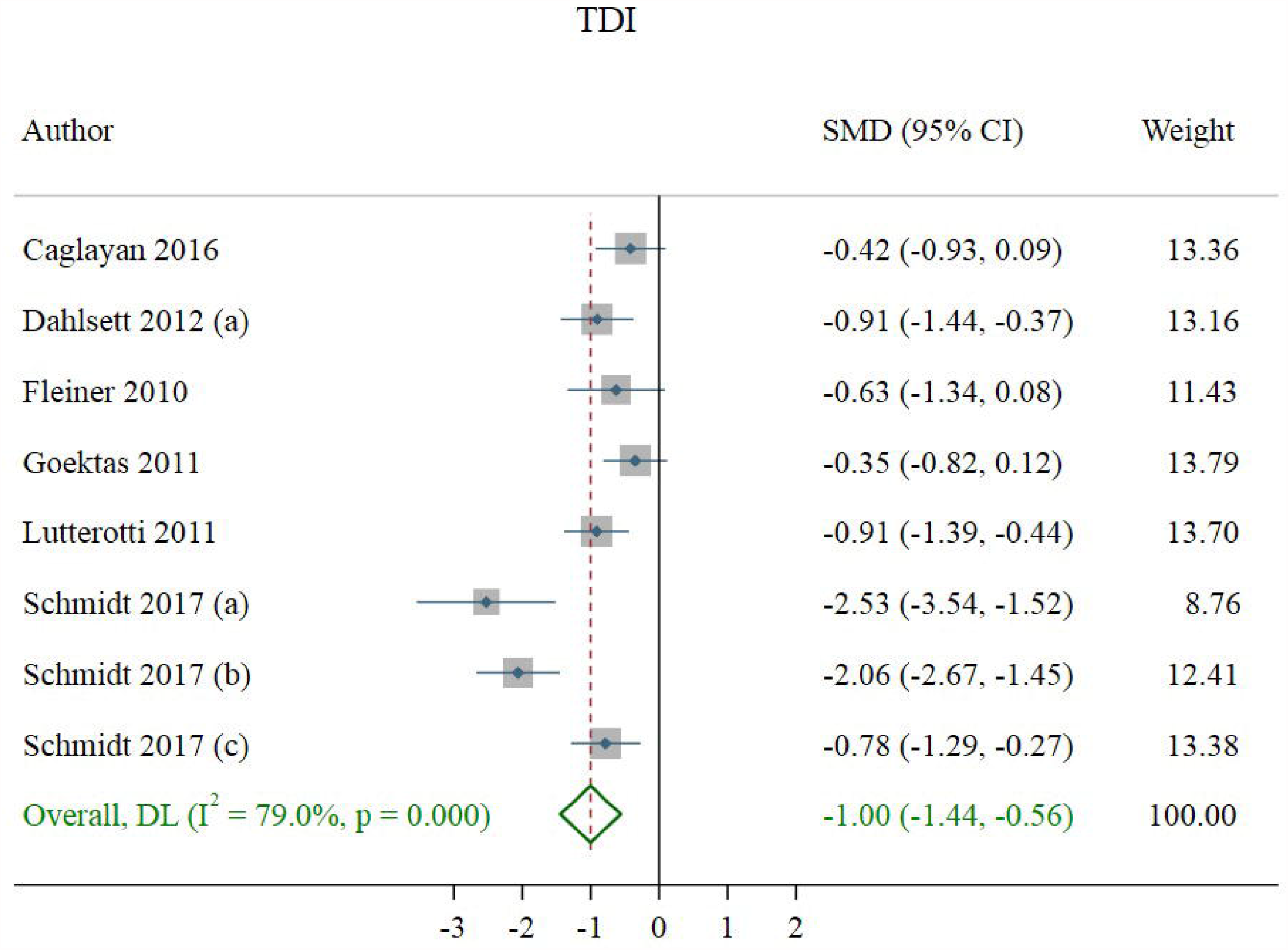
Studies reporting TDI, Threshold, Discrimination and Identification.

**Figure 6.**
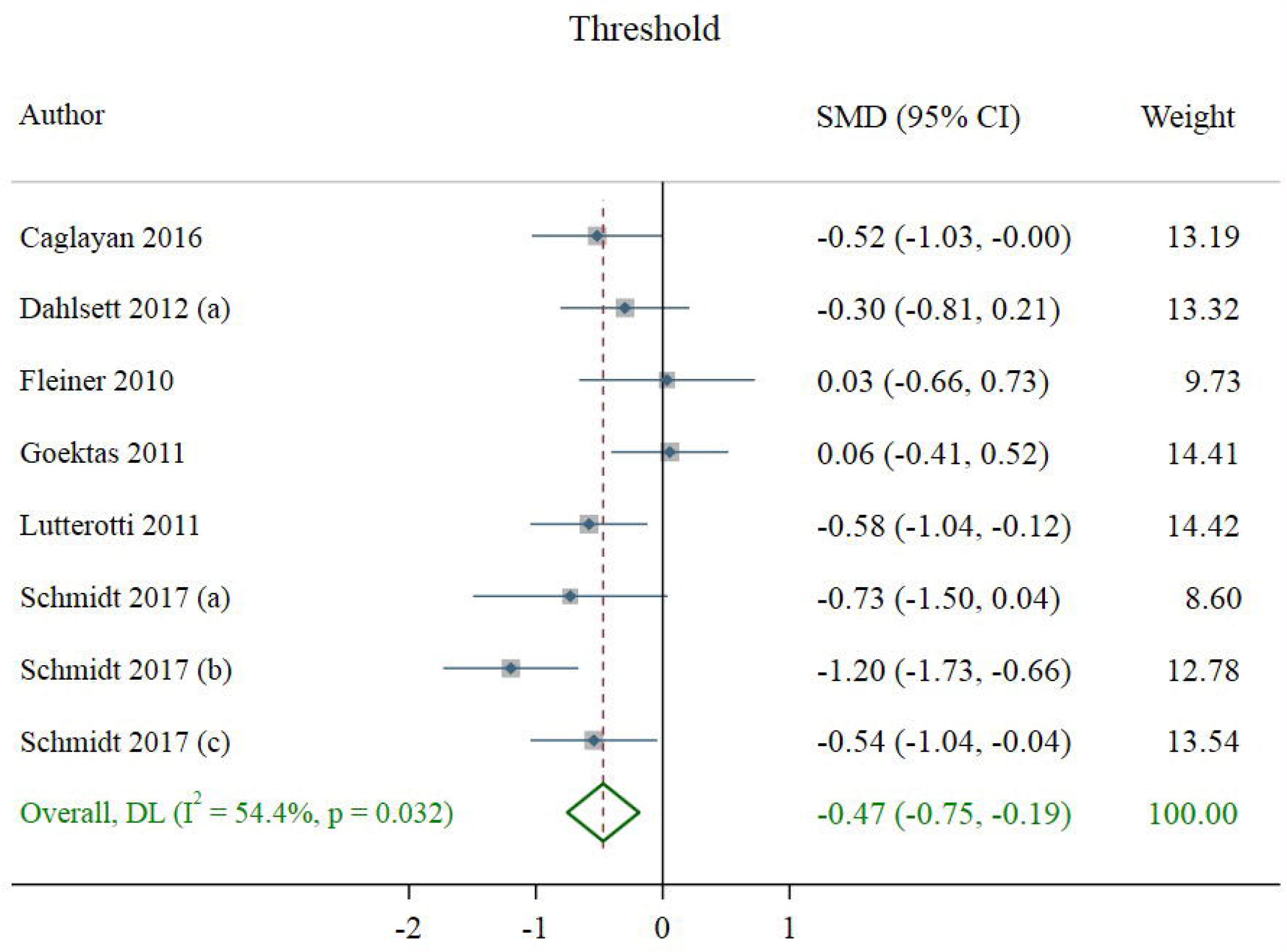
Overall level of Threshold.

**Figure 7.**
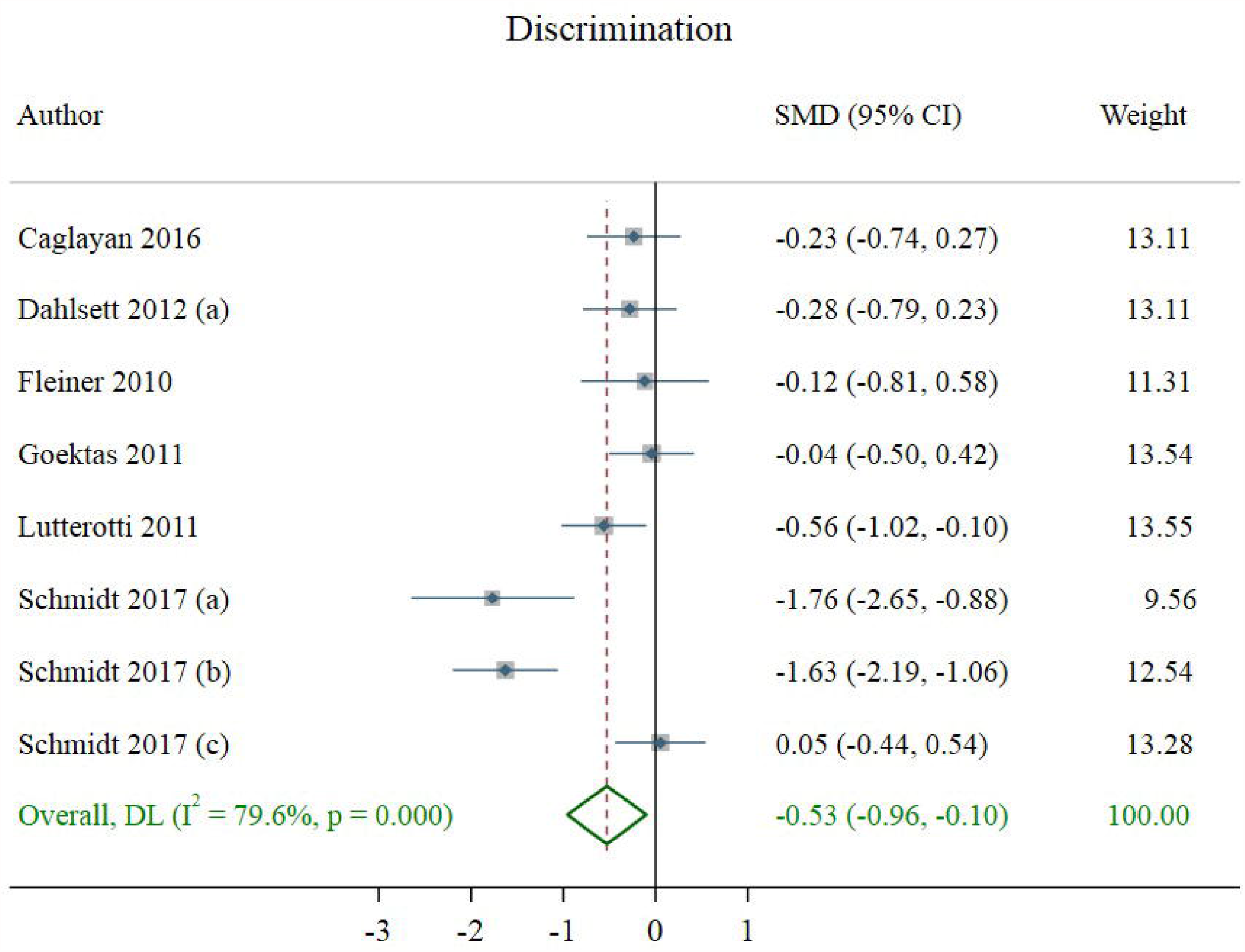
Overall level of Discrimination.

**Figure 8.**
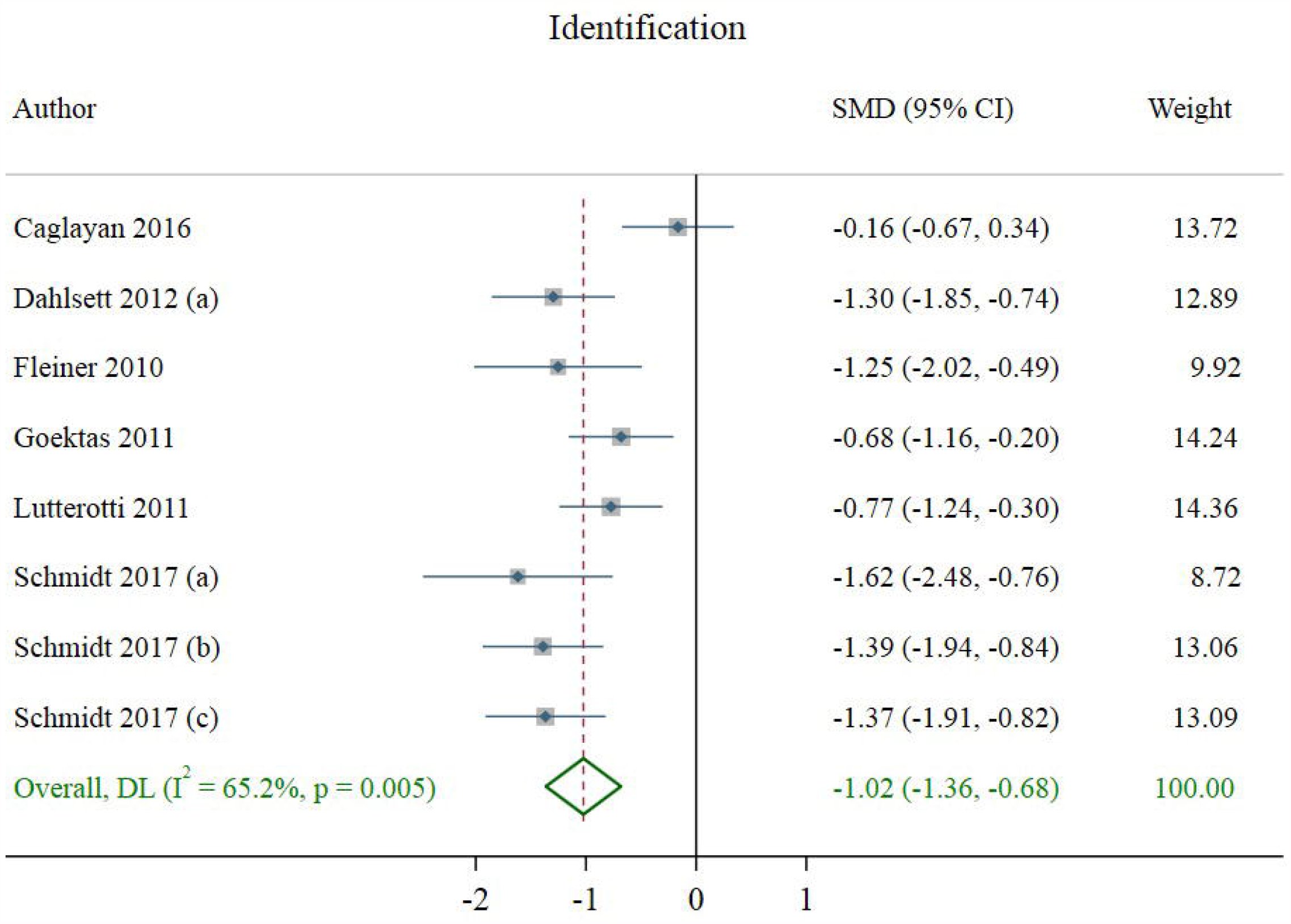
Overall level of Identification.

## Discussion

To our knowledge, this is the first systematic review and meta-analysis on the prevalence of olfactory dysfunction in MS patients. Our results showed that the pooled prevalence of olfactory dysfunction among MS patients was 27.2%. Subgroup analysis showed significantly different prevalence rates between countries. The highest pooled prevalence of olfactory dysfunction among MS patients was seen in Italy with a rate of 69.6%, and the lowest pooled prevalence was observed in Austria at 0.00%. Aside from general olfactory dysfunction, eight studies went into detail and categorized the dysfunction as TDI score, Threshold, Discrimination, and Identification scores. The score is generally reported as in Mean (∓SD) and is utilized to report the findings of the Sniffin’ Sticks-Test and the TDI test. However, some researchers may choose to use the TDI score as an independent screening test and report the three major aspects of olfaction using the aforementioned tool. The overall TDI score in MS patients was lower than that in the control group.

Olfactory dysfunctions are reported to have strong correlations with MS. Not only do MS patients show higher levels of olfactory impairment but they also forfeit this ability as their disease progress. That is why the idea of using olfactory screening tests as a diagnostic and prognostic marker is capturing more and more interest every day. (7,13,14) There are multiple tests to assess one’s ability of olfaction, such tests include but are not limited to the University of Pennsylvania Smell Identification Test (UPSIT), the Sniffin’ Sticks-Test, Odor Stick Identification Test for the Japanese (OSIT-J), Olfactory Evoked Responses Potentials (OERP). Different studies have applied different tests based on their methodology and protocols.(15,16) Nevertheless, the golden standard of diagnosing olfactory dysfunction is the Toyota and Takagi (T&T) Olfactometer. The test utilizes its own specific kit and shall be conducted in a well ventilated and electrically shielded room.(17)

A study conducted in 2012 with a sample size of 153 reported that 11% of MS patients had olfactory dysfunction. This article is unique as it is one of the few studies that reported olfactory dysfunction in the control group as well, at 3%, which is significantly less than that of the MS group.(18)

In a 2020-published Austrian study, Gabriel Bste et. al. evaluated 260 MS patients and found that 27.3% had hyposmia which is much higher than the general population. This study also reported 110 MS patients (42%) were smokers which might be used for future research into the presumably confounding association between smoking and olfactory dysfunction among MS patients.(19)

However, one of the highest rates of olfactory dysfunction among MS patients was reported by F.A. Schmidt et. al., who examined 64 MS patients in 2017 and revealed 57.8% had olfactory dysfunction. It is notable to mention that this was one of the few studies where the Threshold Discrimination Identification (TDI) test was performed as the screening tool.(20)

So far, existing data suggest that MS patients are at an elevated risk of experiencing olfactory dysfunction. Moreover, our subgroup analysis showed that the pooled prevalence of olfactory dysfunction in studies whose mean EDSS was more than 3 was higher compared to other studies. One possible explanation for this is that the intersection of persistent inflammation within the CNS, demyelination of olfactory bulbs, and the burden of plaque in brain areas associated with the olfactory system contribute to the disturbances seen in the olfaction of MS patients.(9,21) As such, pro-inflammatory cytokines which are abundantly found in the CNS of MS patients have been shown to be inversely correlated with olfactory function.(22) Besides, olfactory dysfunction is recognized across an ever-broadening spectrum of demyelinating conditions including MS. Demyelination and MS-plaque formation within the olfactory-related CNS regions are thought to disturb normal olfaction in the same way as it affects other sensory pathways.(8,23) However, unlike previous studies, our results did not show a correlation between olfactory dysfunction an disease duration.(18,24) This might be due to the fact that our analysis reported a high level of heterogeneity.

Olfactory dysfunction is associated with other diseases especially those affecting the CNS as well. A review article by Shin et. al. suggested that aside from MS, neuromyelitis optica, and systemic lupus erythematosus are related to olfactory disorders. Their study also claims that inflammation in the olfactory bulb in animal models results in olfactory disturbances.(2)

The biology behind this cascade can be explained as follows: Data supports the concept that acute, self-limited inflammatory response mediates repair signaling through the NF-κB pathway and contributes to neuro-regeneration in the Olfactory Epithelium (OE).(25) However, once this inflammation gets out of control, more harm is caused than good, which leads to a disruption in the cell-cycle regulation of the OE.(25,26) Such disruptions eventually lead to degeneration of olfactory receptors and its related neural signaling pathways.(27) Moreover, the same pathophysiology is thought to contribute to the ever-growing report of olfactory dysfunction among COVID-19 patients. As the inflammation heightens, the release of pro-inflammatory cytokines interferes with normal olfactory neural signaling pathways, leading to altered states of olfaction.(28) Inflammation seems like the pivotal concept that bridges MS and COVID-19 with olfactory dysfunction, since in both cases the NF-κB pathway signaling seems to play a crucial role. There is accumulating evidence on the role of this signaling pathway in exacerbated inflammation, the development of MS, MS-related sensory-neural disturbances, and COVID-19-driven olfactory dysfunction.(14,25,29,30)

Overall, different aspects of MS pathophysiology seem to be working like building blocks for an altered, disturbed, and dysfunctional olfaction in affected patients.

Our study has some strengths. First, it is the first systematic review evaluating the prevalence of olfactory dysfunction among MS patients. Second, not only did we assess the prevalence of olfactory dysfunction in MS patients but we also estimated its pooled prevalence based on different disease duration and EDSS scores. However, we had some limitations, too. For example, Whether or not an association exist between the type of medication patients receive and the chance of olfactory dysfunction was not assessed.

## Conclusion

The results of this systematic review show that the prevalence of olfactory dysfunction in MS patients is significantly higher than the general population. Also, not only is the overall collective TDI score in MS patients lower than that in the control group, but the level of Threshold, Discrimination, and Identification per se are lower in MS compared with control as well. It also provides us with insight into the importance of routine and systemic checkups in MS patients in an effort to prevent the progression of severe comorbidities.

## Supporting information

Supplementary 1 Table of quality assessment of the included studies using the JBI checklist.

Supplementary 2 Figure of quality assessment of the included studies using the JBI checklist.

## Data Availability

All of the data will be available for secondary analysis in necessary cases from the corresponding author through an email address.

