## Supplementary 1 Table of quality assessment of the included studies using the JBI checklist. for "Olfactory Dysfunction in Patients with Multiple Sclerosis; A Systematic Review and Meta-Analysis"

|  | Q1) Was the sample frame appropriate to address the target population? | Q2) Were study participants sampled in an appropriate way? | Q3) Was the sample size adequate? | Q4) Were the study subjects and the setting described in detail? | Q5) Was the data analysis conducted with sufficient coverage of the identified sample? | Q6) Were valid methods used for the identification of the condition? | Q7) Was the condition measured in a standard, reliable way for all participants? | Q8) Was there appropriate statistical analysis? | Q9) Was the response rate adequate, and if not, was the low response rate managed appropriately? |
| --- | --- | --- | --- | --- | --- | --- | --- | --- | --- |
| Bsteh, 2017 | Yes | Unclear | Yes | Yes | Yes | Yes | Yes | Yes | Unclear |
| Bsteh, 2018 | No | No | No | Yes | Unclear | Yes | Yes | Yes | Unclear |
| Caglayan, 2016 | No | Unclear | No | Yes | No | Yes | Yes | Yes | Unclear |
| Caminiti, 2014 | No | No | No | Yes | Yes | Yes | Yes | Yes | Yes |
| Carotenuto, 2018 | No | No | No | Yes | Yes | Yes | Yes | Yes | Yes |
| Dahlsett, 2012 | No | No | No | No | Unclear | Yes | Yes | Unclear | Unclear |
| Doty,1998 | No | Unclear | No | No | No | Yes | Yes | No | Unclear |
| Fleiner, 2010 | No | No | No | No | Unclear | Yes | Unclear | Unclear | Unclear |
| Goektas, 2011 | No | Yes | No | No | Unclear | Unclear | Yes | Unclear | Unclear |
| Hawkes, 1997 | No | No | No | No | Yes | Unclear | Unclear | Yes | Yes |
| Hawkes, 1998 | Unclear | Unclear | No | No | Unclear | Unclear | Unclear | Unclear | Unclear |
| Holinski, 2014 | No | No | No | No | Unclear | Yes | Yes | Unclear | Unclear |
| Lawrence, 1996 | No | No | No | Yes | No | Yes | Yes | Yes | Unclear |
| Li-Min Li, 2015 | No | Unclear | No | Yes | No | Yes | Yes | Yes | Unclear |
| Li-Min Li, 2018 | No | Unclear | No | No | Yes | Yes | Yes | Yes | Yes |
| Lutterotti, 2011 | No | No | No | No | Unclear | Unclear | Yes | Unclear | Unclear |
| Okadaa, 2020 | No | No | No | Yes | Unclear | Yes | No | Yes | Unclear |
| Schmidt, 2017 | No | Unclear | No | Yes | No | Yes | Yes | Yes | Unclear |
| Silva, 2012 | No | No | Yes | No | Unclear | Yes | Unclear | Unclear | Unclear |
| Uecker, 2017 | No | No | No | Yes | Yes | Yes | Yes | Yes | Yes |
| Zivadinov, 1999 | No | No | No | No | Unclear | Unclear | Unclear | Unclear | Unclear |
| Zorzon, 2000 | No | Unclear | No | Yes | No | Yes | Yes | Yes | Unclear |
