## Supplementary figures and images for "Olfactory Dysfunction in Patients with Multiple Sclerosis; A Systematic Review and Meta-Analysis"

### Supplementary 2 Figure of quality assessment of the included studies using the JBI checklist.

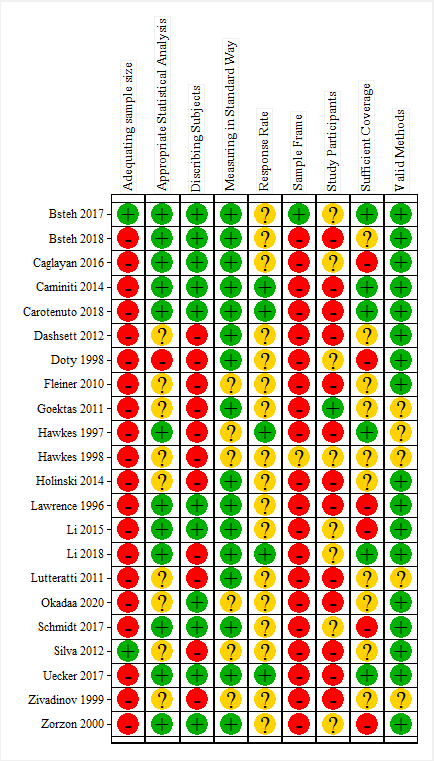
